# Endovascular thrombectomy for the treatment of large ischemic stroke: a systematic review and meta-analysis of randomized control trials

**DOI:** 10.1101/2023.02.27.23286534

**Authors:** Travis J. Atchley, Dagoberto Estevez-Ordonez, Nicholas M.B. Laskay, Borna E. Tabibian, Mark R. Harrigan

**Author notes:** **Corresponding and reprint author:** Mark R. Harrigan, MD, Department of Neurosurgery, University of Alabama at Birmingham, 1060 Faculty Office Tower, 1720 2nd Avenue South, Birmingham, AL 35294. These authors contributed equally to this work (Travis J. Atchley, Dagoberto Estevez-Ordonez).

## Abstract

**Importance:** Endovascular thrombectomy (ET) has previously been reserved for patients with small to medium acute ischemic strokes. Three recent randomized control trials (RCTs) have demonstrated functional benefit and risk profiles for ET in large volume ischemic strokes.

**Objective:** The primary objective of the meta-analysis was to determine the combined benefit of ET in adult patients with large volume acute ischemic strokes and to better determine the risk of adverse events following ET.

**Data Sources:** We systematically searched MEDLINE, EMBASE, SCOPUS, the Cochrane Central Register of Controlled, and Google Scholar for all RCTs published in English language between January 1, 2010, to February 19, 2023.

**Study Selection:** We included only RCTs specifically comparing ET to medical therapy in patients with acute ischemic stroke with large volume infarctions as defined by Alberta Stroke Program Early Computed Tomography Score (ASPECTS) 3-5 or calculated infarct volume of > 50-70mL. Two independent reviewers screened potential studies for full text review and metaanalysis inclusion with conflicts being resolved by consensus or third reviewer.

**Data Extraction and Synthesis:** Data was extracted based on pre-specified variables on study methods and design, participant characteristics, analysis approach, as well as efficacy and safety outcomes. Results were combined using a restricted maximum-likelihood estimation random-effects model. Studies were assessed for potential bias and quality of evidence.

**Main Outcome(s) and Measure(s):** The prespecified primary outcome was an overall ordinal shift across the range of modified Rankin scale scores toward a better outcome at 90 days following either ET or medical management for patients with large volume ischemic strokes.

**Results:** A total of 3044 studies were screened, and 29 underwent full text review. 3 RCTs (1011 patients) were included in the analysis. The pooled random effects model for the primary outcome of mRS improvement favored ET over medical management, generalized odds ratio 1.55 [95% CI 1.25 – 1.91, τ^2^ = 0.01, I^2^ = 42.84%]. There was a trend toward increased risk of symptomatic ICH in the ET group, relative risk 1.85 [95% CI 0.94 – 3.63, τ^2^ = 0.00, I^2^ = 0.00%].

**Conclusions and Relevance:** In patients with large volume ischemic strokes, ET has a clear functional benefit and does not confer increased risk of significant complications compared to medical management alone.

## Introduction

Endovascular thrombectomy (ET) has revolutionized the management for patients with acute large vessel occlusions. Numerous randomized control trials (RCTs) have demonstrated significant benefit in functional outcome (modified Rankin scale (mRS)) compared to medical management alone.^1–4^ Moreover, the benefits of ET have been supported even with increasing time from stroke onset to intervention.^1,5,6^ The vast majority of patients included in these RCTs did not have large-volume ischemic infarcts based upon either computed tomography (CT) perfusion (CTP) studies or via the Alberta Stroke Program Early CT Score (ASPECTS).^2,7^ Current guidelines support ET for large vessel ischemic strokes with ASPECTS ≥ 6, but the role of ET in patients with large-volume infarcts defined as ASPECTS 3-5 or core volumes of > 50-70mL has been less well-defined due to perceived risk of intracranial hemorrhage (ICH) or absence of functional benefit.^8^

In the past year, three multicenter RCTs have been published specifically investigating the benefits of ET in patients with large vessel occlusions and ASPECTS 3-5 while excluding those with very low ASPECTS of 0-2. The RESCUE-Japan LIMIT, ANGEL-ASPECT, and SELECT2 trials were conducted in Japan, China, and an international conglomerate (North America, Europe, Australia, and New Zealand), respectively.^9–11^ These multicenter RCTs have all have demonstrated differing margins of benefit in functional outcome following ET in large-volume ischemic strokes, and they have also reported differing rates of ICH.^9–11^ We sought to perform a systematic review of the literature for any recent RCT that includes large-volume ischemic strokes and to subsequently perform a meta-analysis of these results. This study’s aims were to (1) determine if ET leads to improved outcomes measured via mRS in adult patients with large ischemic strokes when compared to medical management and (2) understand the safety of ET in patients with large ischemic strokes by comparing rates of symptomatic ICH and other adverse events. In analyzing these major trials, we can better understand the true benefit in mRS and risk profile for patients low ASPECTS receiving ET.

## Methods

We conducted a systematic review and meta-analysis according to a prespecified protocol registered at the International Platform of Registered Systematic Review and Meta-analysis Protocols (INPLASY) (registration number INPLASY202320107, **eAppendix 1**). This report adheres to guidelines in the Preferred Reporting Items for Systematic Reviews and MetaAnalyses (PRISMA) 2020 statement.^12^

### Eligibility criteria and information sources

We included RCTs specifically comparing ET to medical management in the setting of an acute ischemic stroke with large core or large volume infarct as defined by ASPECTS 3-5 or calculated core infarct of > 50-70mL. We limited our selection to studies on adult patients (>18 years old). Clinical and observational studies, case series with available abstracts and published as full-scale original articles, brief reports in peer-reviewed academic journals, pilot reports, opinion pieces, theses, conference proceedings, letters, editorials, meta-analysis, reviews, surgical technique papers, case reports, abstracts, presentations, and any non-English language publications without translations were excluded.

### Search strategy

We systematically searched MEDLINE, EMBASE, SCOPUS, the Cochrane Central Register of Controlled Trials (CENTRAL), and Google Scholar. Since the first thrombectomy trials were published nearly two decades ago, we limited our search article published between January 1, 2010, to February 19, 2023.^13^ The search strategy included broad key words to identify studies on patients with ischemic stroke, large vessel occlusion, endovascular treatment, endovascular therapy, and thrombectomy with filters to limit time range (since 2010) and in some instances filters to identify RCTs. The details of the search strategy employed on each database are included in **eAppendix 2**.

### Selection process

After exclusion of duplicates, two reviewers (TJA and BET) independently screened articles for relevance first based on titles and abstracts and subsequently through full-text review to identify articles meeting eligibility criteria. Disagreements between reviewers were resolved in both phases by either consensus or with a third reviewer (DEO). For the selection process, we relied on the Covidence systematic review software (Veritas Health Innovation, Melbourne, Australia; available at www.covidence.org.)^14^

### Data collection

Two investigators (TJA and BET) independently extracted data from each trial meeting eligibility criteria using a standardized excel collection form. Data was extracted according to the protocol and included variables on study characteristics, design, demographic data of enrolled participants, analysis approach (intention-to-treat vs per-protocol), efficacy and safety outcomes of interest as outlined in the protocol, and adherence to reporting guidelines.^15^ We also extracted data for a pre-specified subgroup analysis on the primary outcome using time from stroke onset to randomization, site of arterial occlusion (internal carotid artery (ICA) or middle cerebral artery (MCA), infarct core volume, and ASPECTS.

### Outcomes

The primary outcome for the meta-analysis was an overall ordinal shift across the range of mRS scores toward a better outcome at 90 days. Secondary outcomes included functional independence defined as an mRS score of 0–2 at 90 days and independent ambulation defined as an mRS score of 0–3 at 90 days. Safety outcomes included symptomatic ICH, any ICH, death at 90 days, and need for decompressive HC.

### Risk of bias assessment

Two investigators (NMBL and DEO) with no affiliation to the studies meeting eligibility criteria independently assessed risk of bias for each study using the Cochrane Risk-of-Bias in randomized trials (RoB 2) tool.^16,17^ Disagreements were resolved by consensus or consulting with a third reviewer (TJA).

### Data synthesis

Results of the included trials were combined through meta-analysis to obtain summary estimates of effect sized for each of the pre-specified primary and secondary outcomes. Generalized Odds Ratio (OR) for ordinal shift in mRS scores toward a better outcome were combined by using a random-effect model with restricted maximum-likelihood estimation. All other outcomes were binary, and relative risks (RR) ratios were estimated by combining data from all studies also using a random-effect model with restricted maximum-likelihood estimation. Subgroup analysis was performed only on the primary outcome. Trials with missing data were not used in the meta-analysis. This only occurred in two instances for safety outcomes alone.

We also performed sensitivity analyses by exploring how global effect sizes and p-values were affected by adjusting to the between-study variance parameter τ2 (**eAppendix 3**). Statistical heterogeneity and the magnitude of heterogeneity was assessed using Cochran χ^2^ tests and the *I*^*2*^ statistic, respectively. Publication bias was assessed using the Egger test and visually using funnel plots (**eAppendix 4**).^18^ Statistical analyses were performed using STATA/MP version 17 (StataCorp, College Station, TX).^19^ Alpha was set at 0.05 and all test of significance were 2-sided. To reduce the potential for type I error due to multiple testing, we used Bonferroni corrections to adjust p-values. Data and syntax used for the analysis have been made publicly available through GitHub (https://github.com/estevezdo/Thrombectomy-in-Large-Volume-Strokes-SysRev-Metan).

### Strength of the evidence

We relied on the Grading of Recommendations Assessment, Development, and Evaluation (GRADE) approach to assess the evidence that thrombectomy compared standard care improves outcomes as measured with the mRS.^20^ The combined outcomes were assigned an overall grade of high, moderate, low, or insufficient. This process was completed initially by a single investigator (NMBL) and with consensus after being reviewed by all investigators.

## Results

A total of 3044 studies were screened by title and abstract. Twenty-nine studies underwent full text review for potential meta-analysis inclusion. Of these, only 3 studies met the prespecified inclusion criteria. The most common reasons for study exclusion upon initial screening were the wrong study protocol (ie. not a randomized control trial) and failure to specifically investigate patients with ASPECTS 3-5 or volumes >50-70mL. **Figure 1** demonstrates the PRISMA flow diagram for screening and study inclusion/exclusion. The included studies were assessed for bias via the Cochrane RoB 2 tool.^16,17^ Using the GRADE approach, all evidence from the include studies was assigned a high grade (**eAppendix 5**).^20^

**Figure 1.**
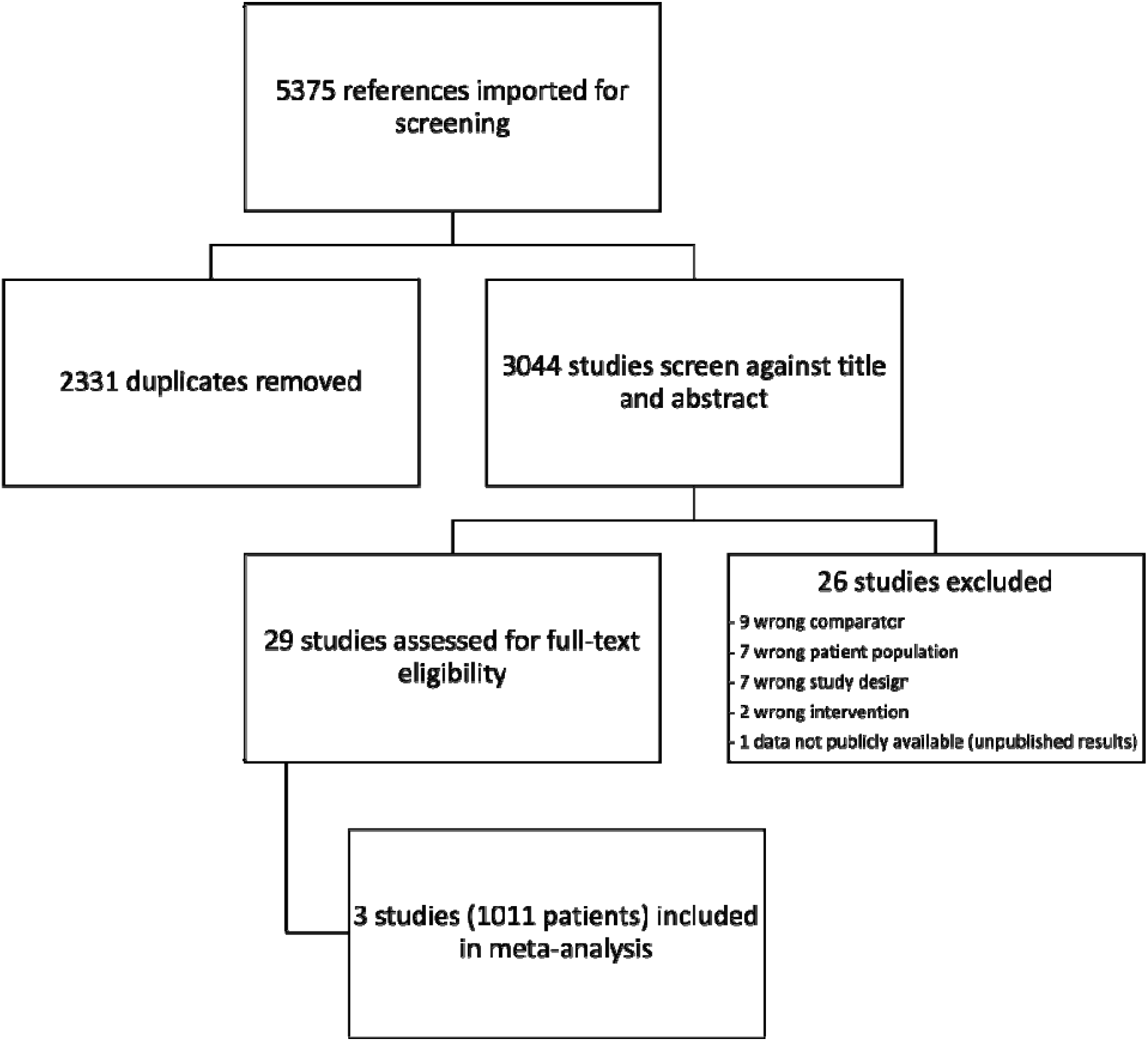
PRISMA Flow Diagram for Study Screening and Inclusion/Exclusion

Among the 3 studies, 1011 patients were included in the meta-analysis. Of these, 510 patient were randomized to ET, and 501 to medical management. Of the 757 patients with available data, 687 (90.7%) had ASPECTS of 3-5. **Table 1** shows the demographic and clinical characteristics of the 3 included trials. The primary outcome of ordinal shift in mRS at 90 days Huo et al., Sarraj et al., and Yoshimura et al. report ORs 1.37 [95% CI 1.11 – 1.69], 1.51 [95% CI 1.20 – 1.89], and 2.43 [95% CI 1.35 – 4.37], respectively. A total of 119/507 (23.5%) and 46/498 (9.2%) patients had functional independence (mRS 0-2) at 90 days in the ET and medical management groups, respectively. A total of 206/507 (40.6%) and 120/498 (24.1%) patients were functional ambulators (mRS 0-3) at 90 days in the ET and medical management groups, respectively. In the ET and medical management groups, 24/507 (4.7%) and 13/498 (2.6%) patients had symptomatic ICH, respectively. The primary, secondary, and safety outcomes of the included trials are shown in **Table 2**.

**Table 1.** Demographic and Clinical Characteristics.

| Characteristic | Huo et al. |  | Sarraj et al. |  | Yoshimura et al. |  |
| --- | --- | --- | --- | --- | --- | --- |
|  | ET | MM | ET | MM | ET | MM |
| Patients randomized | 231 | 225 | 178 | 174 | 101 | 102 |
| Lost to Follow up | 0 | 0 | 1 | 3 | 0 | 0 |
| Analysis intention-to-treat population | 230 | 225 | 178 | 174 | 100 | 102 |
| Analysis per-protocol population | 219 | 222 | 174 | 162 | 94 | 94 |
| Median Age (IQR) – yr | 68 (61-73) | 67 (59-73) | 66 (58-75) | 67 (58-75) | 76.6±10.0* | 75.7±10.2* |
| Median NIHSS score at admission (IQR)† | 16 (13-20) | 15 (12-19) | 19 (15-23) | 19 (15-22) | 22 (18-26) | 22 (17-26) |
| ASPECTs value based on CT§ |  |  |  |  |  |  |
| ASPECTs <3 | 32 (13.9) | 30 (13.3) | 0 | 0 | 5 (5.0) | 3 (2.9) |
| ASPECTs ≥3 | 198 (86.0) | 195 (86.7) | 67 (74.8) | 32 (37.4) | 96 (95.0) | 99 (97.1) |
| Median infarct-core volume (IQR) – mL¶ | 60.5 (29-86) | 63 (31-86) | 81.5 (57-118) | 79 (62-111) | 94 (66-152) | 110 (74-140) |
| Time from stroke onset to randomization <6 hours | 82 (35.7) | 85 (37.8) | 17 (30.1) | 5 (11.4) | 71 (70.3) | 74 (72.6) |
| Time from stroke onset to randomization ≥6 hours | 148 (64.3) | 140 (62.2) | 50 (41.0) | 27 (21.3) | 30 (29.7) | 28 (27.4) |
| Occlusion site – no. (%)‡ |  |  |  |  |  |  |
| ICA | 83 (36.1) | 81 (36.0) | 80 (44.9) | 66 (37.9) | 47 (46.5) | 49 (48.0) |
| M1 segment | 145 (63.0) | 142 (63.1) | 91 (51.1) | 100 (57.5) | 74 (73.3) | 70 (68.6) |
| M2 segment | 2 (0.9) | 2 (0.9) | 7 (3.9) | 8 (4.6) | 0 | 3 (2.9) |

ASPECT = Albert Stroke Program Early Computed Tomography Score; ET = endovascular thrombectomy; ICA = internal carotid artery; IQR = interquartile range; MCA = middle cerebral artery; mL = milliliters; MM = medical management; no. = number; NR = not reported; yr = years
\* Age reported as mean ± standard deviation
† Scores on the National Institutes of Health Stroke Scale (NIHSS), an ordinal scale that is used to evaluate the severity of stroke, range from 0 to 42, with higher scores indicating greater neurologic deficit.
‡ The M1 segment is the first and main segment middle cerebral artery, and the M2 segment is the first branch of the main trunk of the middle cerebral artery.
§ ASPECTS values range from 0 to 10, with lower values indicating larger infarction.
¶ The ischemic-core volume is the volume of irreversibly damaged brain tissue with relative cerebral blood flow of less than 30% of that of the contralateral hemisphere (based on CT perfusion) or an apparent diffusion coefficient of less than $620 \times 10^{-6}$ mm<sup>2</sup> per second (based on MRI).

**Table 2.** Trial Outcomes.

Primary, secondary and safety outcomes of 3 included randomized control trials.
| Table 2. Trial Outcomes |  |  |  |  |  |  |
| --- | --- | --- | --- | --- | --- | --- |
|  | Huo et al. |  | Sarraj et al. |  | Yoshimura et al. |  |
| Outcome | ET<br>(N=230) | MM<br>(N=225) | ET<br>(N=178) | MM<br>(N=174) | ET<br>(N=100) | MM<br>(N=102) |
| Primary Outcome | Median Score on the modified Rankin scale at 90 days (IQR)† |  |  |  | Modified Rankin scale score of 0 to 3 at 90 days – no. (%) |  |
|  | 4 (2-5) | 4 (3-5) | 4 (3-6) | 5 (4-6) | 31 (31.0) | 13 (12.7) |
| Treatment Effect [95% CI]* | 1.37 [1.11 – 1.69] |  | 1.51 [1.20 – 1.89] |  | 2.43 [1.35 – 4.37] |  |
| Secondary Outcomes |  |  |  |  |  |  |
| Functional Independence at 90 days – no. (%)†<br>mRS 0 to 2 | 69 (30.0) | 26 (11.6) | 36/177<br>(20.3) | 12/171<br>(7.0) | 14 (14.0) | 8 (7.8) |
| Independent Ambulation at 90 days – no. (%)†<br>mRS 0 to 3 | 108<br>(47.0) | 75 (33.3) | 67/177<br>(37.9) | 32/171<br>(18.7) | 31 (31.0) | 13 (12.7) |
| Safety Outcomes |  |  |  |  |  |  |
| Symptomatic intracranial hemorrhage within 48hr – no. (%)§¶ | 14 (6.1) | 6 (2.7) | 1 (0.6) | 2 (1.1) | 9 (9.0) | 5 (4.9) |
| Any intracranial hemorrhage within 48hr – no. (%) | 113<br>(49.1) | 39 (17.3) | NR | NR | 58 (58.0) | 32 (31.4) |
| Death within 90 days – no. (%) | 50 (21.7) | 45 (20.0) | 68/177<br>(38.4) | 71/171<br>(41.5) | 18 (18.0) | 24 (23.5) |
| Decompressive hemicraniectomy during hospitalization – no. (%) | 17 (7.4) | 8 (3.6) | NR | NR | 10 (10.0) | 14 (13.7) |

CI = confidence interval; ET = endovascular thrombectomy; hr = hours; IQR = interquartile range; MM = medical management; mRS = modified Rankin scale; no. = number; NR = not reported; yr = years
\* The treatment effect is reported for the primary outcome as a generalized odds ratio with the 95% confidence interval for the ordinal shift in the distribution of scores on the modified Rankin scale toward a better outcome.
† Scores on the modified Rankin scale range from 0 to 6, with higher scores indicating greater disability.
‡ Data on the NIHSS score were missing for three patients in medical-management group.
§ Hou et al. defined symptomatic intracranial hemorrhage according to the Heidelberg bleeding classification (an increase in the NIHSS score of ≥4 points or an increase in the score for an NIHSS subcategory of ≥2 points with any intracranial hemorrhage on imaging).<sup>26</sup>
¶ Sarraj et al. and Yoshimura et al. defined symptomatic intracranial hemorrhage according to Safe Implementation of Thrombolysis in Stroke–Monitoring Study criteria (parenchymal hemorrhage type 2 or remote parenchymal hemorrhage associated with an increase of 4 or more points in the NIHSS score at the 24-hour follow-up).<sup>25</sup>

The generalized OR for ordinal improvement in mRS from the combined studies was 1.55 [95% CI 1.25 – 1.91, τ^2^ = 0.01, I^2^ = 42.84%]. For the outcome of functional independence (mRS 0–2) at 90 days, the combined results favored ET over medical management with RR 2.53 [95% CI 1.84 – 3.47, τ^2^ = 0.00, I^2^ = 0.00%]. For the secondary outcome of independent ambulation (mRS 0–3) at 90 days, the combined study results also favored ET over medical management with RR 1.78 [95% CI 1.29 – 2.46, τ^2^ = 0.05, I^2^ = 56.80%]. The forest plots of these random-effects models are shown in **Figure 1**. There was a trend toward increased risk of symptomatic ICH with the ET group, RR 1.85 [95% CI 0.94 – 3.63, τ^2^ = 0.00, I^2^ = 0.00%]. There was an increased risk of any ICH in the ET group in the Yoshimura et al. and Huo et al. studies, RR 2.30 [95% CI 1.51 – 3.49, τ^2^ = 0.06, I^2^ = 70.22%]. There were no significant differences between death at 90 days and need for decompressive hemicraniectomy (HC) for the two treatment arms of the combined studies with RR 0.95 [95% CI 0.78 – 1.16, τ^2^ = 0.00, I^2^ = 0.00%] and RR 1.22 [95% CI 0.44 – 3.40, τ^2^ = 0.39, I^2^ = 70.28%], respectively. The forest plots of the random-effects models for the safety outcomes are shown in **Figure 2**. The forest plots for the subgroup random effects models for time from stroke onset to randomization (<6 or ≥6 hours), location of occlusion (ICA or MCA), core volume (<70 or ≥70mL), and admission ASPECTS (<3 or ≥3) are shown in **Figure 3**.

**Figure 2.**
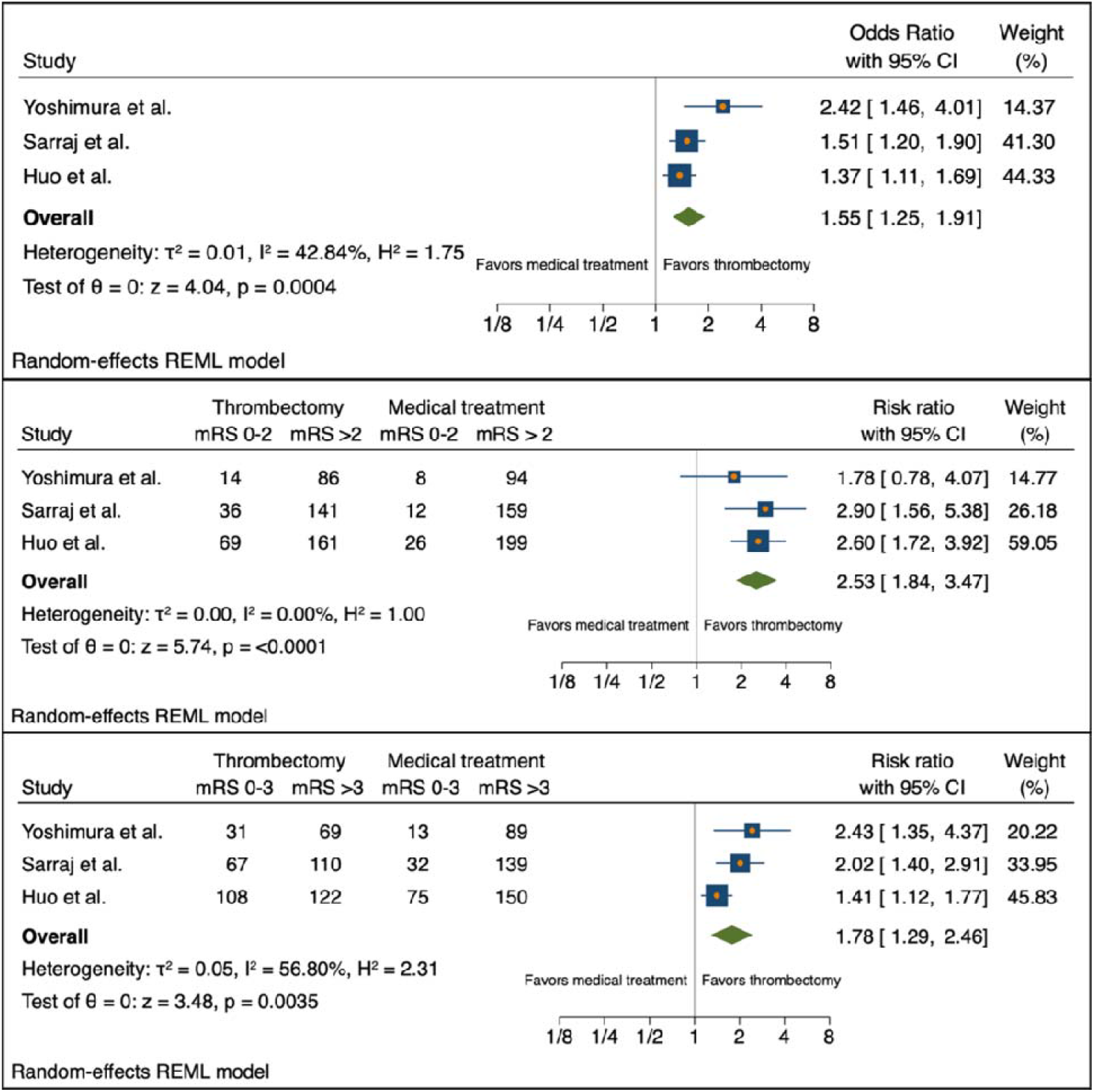
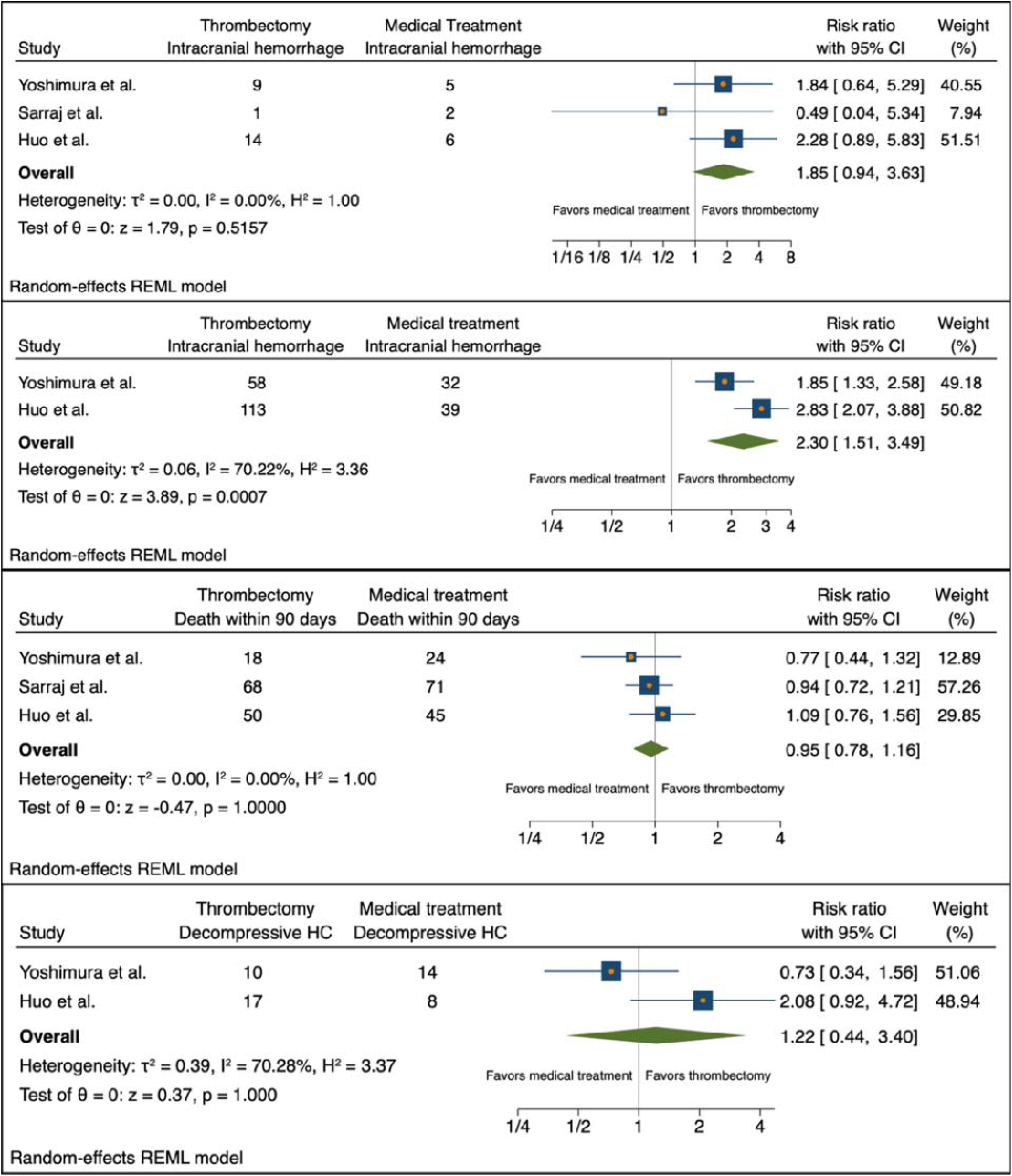
Forest plots for random-effects models for primary and secondary outcomes. CI = confidence interval; HC = hemicraniectomy; mRS = modified Ranking scale; REML = restricted maximum likelihood.

**Figure 3.**
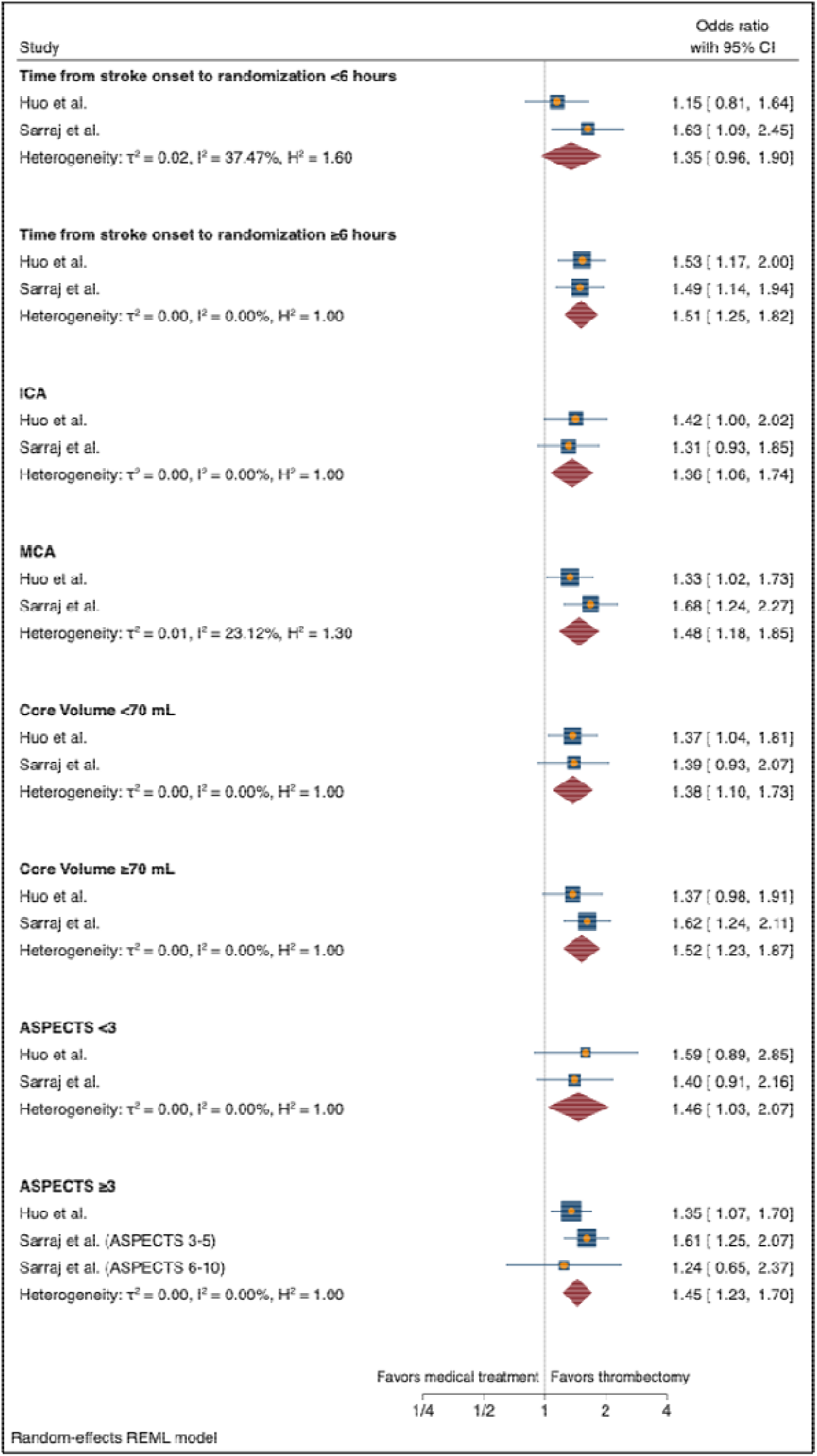
Subgroup analysis forest plots of random effects models. ASPECTS = Albert Stroke Program Early Computed Tomography Score; CI = confidence interval; ICA = internal carotid artery; MCA = middle cerebral artery; mL = milliliters; REML = restricted maximum likelihood.

## Discussion

This combined analysis of the RESCUE-Japan LIMIT, SELECT2, and ANGEL-ASPECT trials, which enrolled patients with large core anterior circulation ischemic strokes within 24 hours of last seen well and randomly assigned to ET or medical care only, confirms the benefit of thrombectomy in terms of 90-day mRS improvements.^9–11^ The benefit of ET was also confirmed in terms of proportions of 90-day functional independence and independent ambulation. Regarding safety of ET in this population, analysis of data from all three trials found no significant difference in proportions of symptomatic intracerebral hemorrhage and mortality.

The MR CLEAN, ESCAPE, REVASCAT, SWIFT PRIME, and EXTEND-IA trials, all published in 2015, provided the first high-level medical evidence supporting the use of ET in selected patients with anterior circulation large-artery occlusion ischemic stroke.^3,4,21–23^ These trials were designed as explanatory trials; the investigators sought to maximize the chance of detecting a benefit with ET by excluding patient subgroups, such as those with large cores, for whom benefit seemed less certain. Large core infarctions have been excluded largely out of concern that post-thrombectomy brain hemorrhage risk would be significant or that the functional benefit would be minimal. However, a meta-analysis of observational studies found favorable results after thrombectomy for patients with acute ischemic stroke and large ischemic cores, typically defined as an ASPECTS score of <6, without significant increased risk of hemorrhage.^24^ The three trials included in the present meta-analysis are the first multicenter RCT of ET for patients with large core infarctions or ASPECTS 3-5.

This study detected a significant benefit with ET for two subgroups that were not found to be significant in the individual studies. Whereas patients with MCA occlusions benefited from thrombectomy in the SELECT2, and ANGEL-ASPECT trials, patients with ICA occlusions were not found to benefit in the individual trials. In the combined analysis, patients with ICA occlusions were found to benefit; this likely reflects the greater severity of ICA occlusions versus MCA occlusions, as well as a greater clot burden and number of needed thrombectomy passes, and therefore a slimmer margin of benefit with ET. Similarly, neither the SELECT2 nor the ANGEL-ASPECT trial found a benefit with ET for patients ASPECTS <3 while the combined analysis did find a benefit for this small subgroup. This likely reflects the greater severity of very low ASPECTS strokes, possibly reduced collateral circulation to the affected territory, and a narrower margin of benefit with ET.

Limitations of this study include limitations of the trials themselves and limitations of the meta-analysis. Regarding the trials themselves, the RESCUE-Japan LIMIT relied only on ASPECTS for identification of patients with a large core, whereas the SELECT2 and ANGEL-ASPECT trials also employed perfusion imaging, which may have improved accuracy in patient selection in those trials compared to the RESCUE-Japan LIMIT trial. Enrollment in the SELECT2 and ANGEL-ASPECT trials was halted early because of efficacy, raising the possibility of overestimation of treatment effect. This also limited the statistical power of the subgroup analyses. Although the RESCUE-Japan LIMIT and ANGEL-ASPECT trials enrolled only Japanese and Chinese patients, raising questions about the generalizability of the findings to non-Asian populations, the SELECT2 trial was largely comprised of non-Asian study subjects and provides evidence that benefit of ET for large core infarctions extends across ethnic groups.

All 3 RCTs incorporate a radiological-symptomatic classification scheme to determine symptomatic ICH. SELECT2 and RESUE-Japan LIMIT trials defined symptomatic ICH according to Safe Implementation of Thrombolysis in Stroke–Monitoring Study criteria (SITS-MOST), while ANGEL-ASPECT defined this outcome by the Heidelberg bleeding classification.^25,26^ However, the SITS-MOST symptomatic ICH criteria was developed before ET devices were being utilized, which may have limited the SELECT2 and RESUE-Japan LIMIT trials from capturing all hemorrhagic complication effects from device recanalization. Additionally, symptoms associated with a hemorrhagic complication are variable and are not clearly represented by just an increase in NIHSS ≥4 shortly after hemorrhage. Thus, the true severity of hemorrhagic complications in the context of a large ischemic stroke also may have been underestimated in these studies.

This meta-analysis was limited by the fact that RESCUE-Japan LIMIT had a different primary endpoint than the other two trials, which constrained combined subgroup analyses only to the SELECT2 and ANGEL-ASPECT trials. This meta-analysis was also limited by the pooled data available from the included studies; a meta-analysis including individual patient data would permit a more granular and precise assessment of treatment effect.

## Conclusion

The findings of this study strengthen the evidence for benefit of ET over medical management for patients with acute ischemic stroke and large core infarctions. There was no increased risk of symptomatic ICH or other adverse events in this group compared to medical management. ET should not be withheld for patients with ASPECTS 3-5 or large ischemic cores based on perfusion imaging.

## Data Availability

Data and syntax used for the analysis have been made publicly available through GitHub

https://github.com/estevezdo/Thrombectomy-in-Large-Volume-Strokes-SysRev-Metan

