## Supplemental Materials and Data for "Endovascular thrombectomy for the treatment of large ischemic stroke: a systematic review and meta-analysis of randomized control trials"

**eAppendix 1**. Study design and protocol as registered with International Platform of Registered Systematic Review and Meta-analysis Protocols (INPLASY) (registration number INPLASY202320107.

**Endovascular thrombectomy for the treatment of large ischemic stroke: a systematic review and meta-analysis of randomized control trials**

**Version 2.0**

**Amendments –**

**Changes made pre-analysis.**

**Added ASPECTS to subgroup analysis.**

**Added search strategies/queries to the protocol.**

**Added GRADE to confidence in cumulative evidence section.**

**Corrections made to selection and data collection processes.**

**Corrected an error/inconsistency in data synthesis portion by eliminating last sentence.**

**Title Page**

Authors:

Travis J. Atchley, MD

Dagoberto Estevez-Ordonez, MD

Nicholas M.B. Laskay, MD

Borna E. Tabibian, MD

Mark R. Harrigan, MD

Study Contributions:

TJA, NMBL, DEO, BET, MRH oversight of design, review, and supervision

TJA, NMBL, DEO, MRH study design, data analysis, guarantors of review

MRH oversight of design

TJA, BET independent screening

TJA, NMBL, DEO data extraction

DEO, NMBL statistical analysis

TJA, NMBL, DEO, BET, MRH manuscript writing and review

Corresponding Author:

Mark R. Harrigan, MD

Professor

Department of Neurosurgery, University of Alabama at Birmingham, Alabama

Funding/Support:

This project is supported in part by the National Institute of Neurological Disorders and Stroke of the National Institutes of Health under award number R25NS079188 (DEO). The content is solely the responsibility of the authors and does not necessarily represent the official views of the National Institutes of Health. DEO is also a Cornwall Clinical Scholar supported by the University of Alabama at Birmingham.

**Introduction**

Rationale:

Endovascular therapy (ET) has revolutionized the management for patients with acute large vessel occlusions (LVO). Numerous randomized control trials (RCTs) have demonstrated significant benefit in functional outcome (modified Rankin scale (mRS)) compared to medical management alone.^1–4^ Moreover, the benefits of endovascular therapy have been supported even with increasing time from stroke onset to intervention.^1,5,6^ The vast majority of patients included in these RCTs did not have large-volume ischemic infarcts based upon either computed tomography (CT) perfusion (CTP) studies or via the Alberta Stroke Program Early CT Score (ASPECTS).^2,7^ Current guidelines support ET for large vessel ischemic strokes with ASPECTS ≥ 6, but the role of ET in patients with large-volume infarcts defined as ASPECTS 3-5 has been less well-defined due to perceived risk of intracranial hemorrhage (ICH) or absence of functional benefit.^8^

In the past year, three multicenter RCTs have been published specifically investigating the benefits of ET in patients with LVO and ASPECTS 3-5. The RESCUE-Japan LIMIT, ANGEL-ASPECT, and SELECT2 trials were conducted in Japan, China, and an international conglomerate (North America, Europe, Australia, and New Zealand), respectively.^9–11^ These multicenter RCTs have all have demonstrated differing margins of benefit in functional outcome following ET in large-volume ischemic strokes, and they have also reported differing rates of ICH.^9–11^ We sought to perform a systematic review of the literature for any recent RCT that includes large-volume ischemic strokes and to subsequently perform a meta-analysis of these results. In analyzing these major trials, we can better understand the true benefit in mRS and risk profile for patients with large-volume strokes receiving ET.

Aims:

1. Efficacy: Determine if thrombectomy leads to improved outcomes measured through the modified Rankin scale (mRs) in adult patients with large ischemic strokes when compared to medical management.
2. Safety: Determine the safety of thrombectomy in patients with large ischemic strokes by comparing rates of symptomatic intracranial hemorrhage, death, neurologic worsening (increase of ≥4 points in the NIHSS score within 24 hours after presentation), and procedural complications.

Research Question

Patients:

- Adult patients (>18) with ischemic stroke within 24-hour onset
- Pre-stroke modified Rankin scale (mRs) of 0 or 1
- Large infarct defined as meeting either of the following criteria:
  - Alberta Stroke Program Early Computed Tomography Score (ASPECTS) value of 3 to 5
  - An estimated ischemic-core volume of 50 mL or greater

Intervention: Endovascular therapy (thrombectomy)

Comparator: Medical management

Outcomes:

1. Ordinal shift across the range of modified Rankin scale scores toward a better outcome at 90 days
2. Functional independence defined as a score on the modified Rankin scale of 0 to 2 at 90 days
3. Independent ambulation (a score on the modified Rankin scale of 0 to 3) at 90 days

Other outcomes: Symptomatic ICH, death, neurologic worsening (increase of ≥4 points in the NIHSS score within 24 hours after presentation), and procedural complications.

**Methods/Approach**

Study Design:

Systematic review of the literature and meta-analysis using aggregate-level data and adhering to the PRISMA checklist guideline ^12,13^

Inclusion/Exclusion Criteria:

*Inclusion:*

Study inclusion criteria:

- Randomized clinical trials published since 2010.

Patient/participant inclusion criteria:

- Adult patients (>18) with ischemic stroke within 24-hour onset
- Pre-stroke modified Rankin scale (mRs) of 0 or 1.
- Large infarct defined as meeting either of the following criteria:
  - Alberta Stroke Program Early Computed Tomography Score (ASPECTS) value of 3 to 5
  - An estimated ischemic-core volume of 50 mL or greater

*Exclusions:*

Study exclusion criteria:

- Clinical and observational studies, case series with available abstracts and published as full-scale original articles, brief reports in peer-reviewed academic journals, pilot reports, opinion pieces, theses, conference proceedings, letters, editorials, meta-analysis, reviews, surgical technique papers, case reports, abstracts, presentations, and any non-English language publications without translations.

Information sources:

Medline, Embase, Scopus, Cochrane Central, Google Scholar, and PubMed

Search Strategy:

*Pubmed*

((ischemic stroke[Title/Abstract]) OR (large vessel occlusion[Title/Abstract])) AND ((endovascular treatment[Title/Abstract]) OR (endovascular therapy[Title/Abstract]) OR (thrombectomy[Title/Abstract])) AND (((randomized[Title/Abstract]) OR (randomised[Title/Abstract])) AND ((trial[Title/Abstract]) OR (study[Title/Abstract])))

Filters:

- Years 2010-present

((ischemic strokes[Title/Abstract]) OR (large vessel occlusion[Title/Abstract])) AND ((endovascular treatment[Title/Abstract]) OR (endovascular therapy[Title/Abstract]) OR (thrombectomy[Title/Abstract])) AND (((randomized[Title/Abstract]) OR (randomised[Title/Abstract])) AND ((trial[Title/Abstract]) OR (study[Title/Abstract])))

Filters:

- Years 2010-2023

*Embase*

('ischemic stroke':ti,ab OR 'large vessel occlusion':ti,ab) AND ('endovascular treatment':ti,ab OR 'endovascular therapy':ti,ab) AND (randomised:ti,ab OR randomized:ti,ab OR 'randomized controlled trial'/exp)

*Cochrane Central*

((ischemic stroke) OR (large vessel occlusion)) AND ((endovascular treatment) OR (endovascular therapy) OR (thrombectomy)) AND (((randomized) OR (randomised)) AND ((trial) OR (study))) in Title Abstract Keyword

Filters:

- Years 2010-2023
- English Language

*Scopus*

TITLE-ABS-KEY ( ( ( ischemic AND stroke ) OR ( large AND vessel AND occlusion ) ) AND ( ( endovascular AND treatment ) OR ( endovascular AND therapy ) OR ( thrombectomy ) ) AND ( ( ( randomized ) OR ( randomised ) ) AND ( ( trial ) OR ( study ) ) ) ) AND ( LIMIT-TO ( PUBYEAR , 2023 ) OR LIMIT-TO ( PUBYEAR , 2022 ) OR LIMIT-TO ( PUBYEAR , 2021 ) OR LIMIT-TO ( PUBYEAR , 2020 ) OR LIMIT-TO ( PUBYEAR , 2019 ) OR LIMIT-TO ( PUBYEAR , 2018 ) OR LIMIT-TO ( PUBYEAR , 2017 ) OR LIMIT-TO ( PUBYEAR , 2016 ) OR LIMIT-TO ( PUBYEAR , 2015 ) OR LIMIT-TO ( PUBYEAR , 2014 ) OR LIMIT-TO ( PUBYEAR , 2013 ) OR LIMIT-TO ( PUBYEAR , 2012 ) OR LIMIT-TO ( PUBYEAR , 2011 ) OR LIMIT-TO ( PUBYEAR , 2010 ) ) AND ( LIMIT-TO ( LANGUAGE , "English" ) ) AND ( LIMIT-TO ( SRCTYPE , "j" ) ) AND ( LIMIT-TO ( DOCTYPE , "ar" ) )

*Google scholar*

("randomized clinical trial" OR "randomised clinical trial") AND ("endovascular therapy" OR "endovascular treatment" OR "thrombectomy") AND ("ischemic stroke" OR "large vessel occlusion") AND ("large core" OR "large volume")

Filters:

- Years 2010-2023

Study records:

- Data Management

Two databases will be created for this study. One database will be for selected studies (study design, sample size, year of publications, PMID, database, etc.) and the second database will be for data extraction with preselected variables for meta-analysis (see Data Items below).

- Selection Process

Two independent reviewers will assess remaining articles for relevance first based on titles and abstracts, and then will assess full-text articles for eligibility. Disagreements between reviewers will be resolved in both phases by either consensus or by a third reviewer.

- Data Collection Process

Each selected study will be distributed to two individuals for data extraction in duplicate using an excel database with preselected variables (see data items below). We anticipate no effort needed to contact authors of selected studies to obtain patient level data.

Data Items for extraction:

Study: (First author name followed by *et al.*)

Year of publication

Effect size for each pre-defined outcome variable

Upper limit CI for each pre-defined outcome variable

Lower limit CI for each pre-defined outcome variable

Study size (number of patients enrolled in the trial and on each arm)

Standard Error

Demographic and patient enrollment characteristics

Metadata:

- Journal name where study was published.
- Year of publication
- Enrollment criteria
- Analysis approach: intention-to-treat vs per-protocol.
- Adherence to CONSORT
- Potential sources of bias

Outcomes and Prioritization:

- Primary outcome:
  - Ordinal shift across the range of modified Rankin scale scores toward a better outcome at 90 days
- Secondary outcomes:
  - Functional independence defined as a score on the modified Rankin scale of 0 to 2 at 90 days.
  - Independent ambulation (a score on the modified Rankin scale of 0 to 3) at 90 days
  - Safety outcomes: Symptomatic intracerebral hemorrhage, any intracerebral hemorrhage, death at 90 days, and need for decompressive hemicraniectomy.

Planned Subgroup Analysis:

- All subgroups with equivocal ORs (95% CI crosses 1.0) in the individual studies will be combined in attempt to better determine the potential impact these factors may have on primary and secondary outcomes when larger numbers are present.
- These subgroups will include:
  - Age, time from LKW, use of IV thrombolytics, size of ischemic core volume, cervical ICA occlusion, and ASPECTS.

Outcome Rationale:

The aforementioned outcomes were the primary and secondary outcomes for recently published RCTs examining the role that endovascular therapy has for acute ischemic stroke with large infarcts.^9,10^ The mRS is a validated, clinician-reported measure of global neurologic disability and is widely applied in the stroke literature to evaluate stroke patient outcomes and as an endpoint in RCTs.^14^ The tool comprises 7 grades (0-6) of stroke severity ranging from 0 or “no symptoms at all” to 5 “severe disability” and 6 “death.”^15^ Thus, an ordinal shift across the range of mRS, our primary outcome measure, is the most well validated measure of the degree to which endovascular therapy in patients with large core infarcts affects neurological disability versus no endovascular therapy. Secondary outcomes, such as functional independence, independent ambulation, and safety outcomes were all secondary outcomes of the aforementioned recent RCTs.^9–11^ Functional independence has been measured in the most recent RCTs and determined to be a score of 0 to 2 on the mRS at an endpoint of 90 days at follow up.^9–11^ Independent ambulation has been determined to be a score of 0 to 3 on the mRS.^16^ Late endovascular therapy, especially in patients with a large core ischemic stroke, is has been debated and determination of the appropriate candidate for endovascular therapy must be weighed against the increasing risk of ICH into this area of dead brain.^17^ Thus, safety outcomes such as symptomatic ICH, death, neurological worsening, and procedural complications are important to evaluate in the setting of endovascular therapy.

Risk of bias of individual studies:

Risk of bias will be determined at the study level:

- Should there be randomized control trials, we plan on employing the Risk of Bias in randomized trials (RoB 2) tool.^18^
- In any study competing interests in each study will be noted if any author had ties to industry, particularly those funded by an industry sponsor, have the potential for bias in favor of the sponsor’s product or if such information was not disclosed.
- Studies will be assessed on quality based on compliance to EQUATOR network guidelines.^19^

We plan on using a funnel plot using Egger tests to assess the for publication bias.^20^

Data synthesis:

We expect variability in patient selection among the RCTs. Therefore, we plan on using a random‐effect model with restricted maximum-likelihood estimation to perform.^21^ We plan on using an inconsistency index (I2) to assess for heterogeneity. ^22^

Confidence in cumulative evidence:

Studies will be assessed on quality based on compliance to EQUATOR network guidelines (CONSORT).^19^ We will also use the Grading of Recommendations Assessment, Development, and Evaluation (GRADE) approach to assess the evidence that thrombectomy compared standard care improves outcomes as measured through mRS.

18.	Risk of bias tools. https://www.riskofbias.info/.

19.	EQUATOR Network | Enhancing the QUAlity and Transparency Of Health Research. https://www.equator-network.org/.

20.	Egger, M., Davey Smith, G., Schneider, M. & Minder, C. Bias in meta-analysis detected by a simple, graphical test. *BMJ* **315**, 629–634 (1997).

21.	DerSimonian, R. & Laird, N. Meta-analysis in clinical trials. *Control. Clin. Trials* **7**, 177–188 (1986).

22.	Higgins, J. P. T., Thompson, S. G., Deeks, J. J. & Altman, D. G. Measuring inconsistency in meta-analyses. *BMJ* **327**, 557–560 (2003).

**eAppendix 2**. Detailed search strategy employed for each database.

Information sources:

Medline, Embase, Scopus, Cochrane Central, Google Scholar, and PubMed

Search Strategy:

*Pubmed*

((ischemic stroke[Title/Abstract]) OR (large vessel occlusion[Title/Abstract])) AND ((endovascular treatment[Title/Abstract]) OR (endovascular therapy[Title/Abstract]) OR (thrombectomy[Title/Abstract])) AND (((randomized[Title/Abstract]) OR (randomised[Title/Abstract])) AND ((trial[Title/Abstract]) OR (study[Title/Abstract])))

Filters:

- Years 2010-present

((ischemic strokes[Title/Abstract]) OR (large vessel occlusion[Title/Abstract])) AND ((endovascular treatment[Title/Abstract]) OR (endovascular therapy[Title/Abstract]) OR (thrombectomy[Title/Abstract])) AND (((randomized[Title/Abstract]) OR (randomised[Title/Abstract])) AND ((trial[Title/Abstract]) OR (study[Title/Abstract])))

Filters:

- Years 2010-2023

*Embase*

('ischemic stroke':ti,ab OR 'large vessel occlusion':ti,ab) AND ('endovascular treatment':ti,ab OR 'endovascular therapy':ti,ab) AND (randomised:ti,ab OR randomized:ti,ab OR 'randomized controlled trial'/exp)

*Cochrane Central*

((ischemic stroke) OR (large vessel occlusion)) AND ((endovascular treatment) OR (endovascular therapy) OR (thrombectomy)) AND (((randomized) OR (randomised)) AND ((trial) OR (study))) in Title Abstract Keyword

Filters:

- Years 2010-2023
- English Language

*Scopus*

TITLE-ABS-KEY ( ( ( ischemic AND stroke ) OR ( large AND vessel AND occlusion ) ) AND ( ( endovascular AND treatment ) OR ( endovascular AND therapy ) OR ( thrombectomy ) ) AND ( ( ( randomized ) OR ( randomised ) ) AND ( ( trial ) OR ( study ) ) ) ) AND ( LIMIT-TO ( PUBYEAR , 2023 ) OR LIMIT-TO ( PUBYEAR , 2022 ) OR LIMIT-TO ( PUBYEAR , 2021 ) OR LIMIT-TO ( PUBYEAR , 2020 ) OR LIMIT-TO ( PUBYEAR , 2019 ) OR LIMIT-TO ( PUBYEAR , 2018 ) OR LIMIT-TO ( PUBYEAR , 2017 ) OR LIMIT-TO ( PUBYEAR , 2016 ) OR LIMIT-TO ( PUBYEAR , 2015 ) OR LIMIT-TO ( PUBYEAR , 2014 ) OR LIMIT-TO ( PUBYEAR , 2013 ) OR LIMIT-TO ( PUBYEAR , 2012 ) OR LIMIT-TO ( PUBYEAR , 2011 ) OR LIMIT-TO ( PUBYEAR , 2010 ) ) AND ( LIMIT-TO ( LANGUAGE , "English" ) ) AND ( LIMIT-TO ( SRCTYPE , "j" ) ) AND ( LIMIT-TO ( DOCTYPE , "ar" ) )

*Google scholar*

("randomized clinical trial" OR "randomised clinical trial") AND ("endovascular therapy" OR "endovascular treatment" OR "thrombectomy") AND ("ischemic stroke" OR "large vessel occlusion") AND ("large core" OR "large volume")

Filters:

- Years 2010-2023

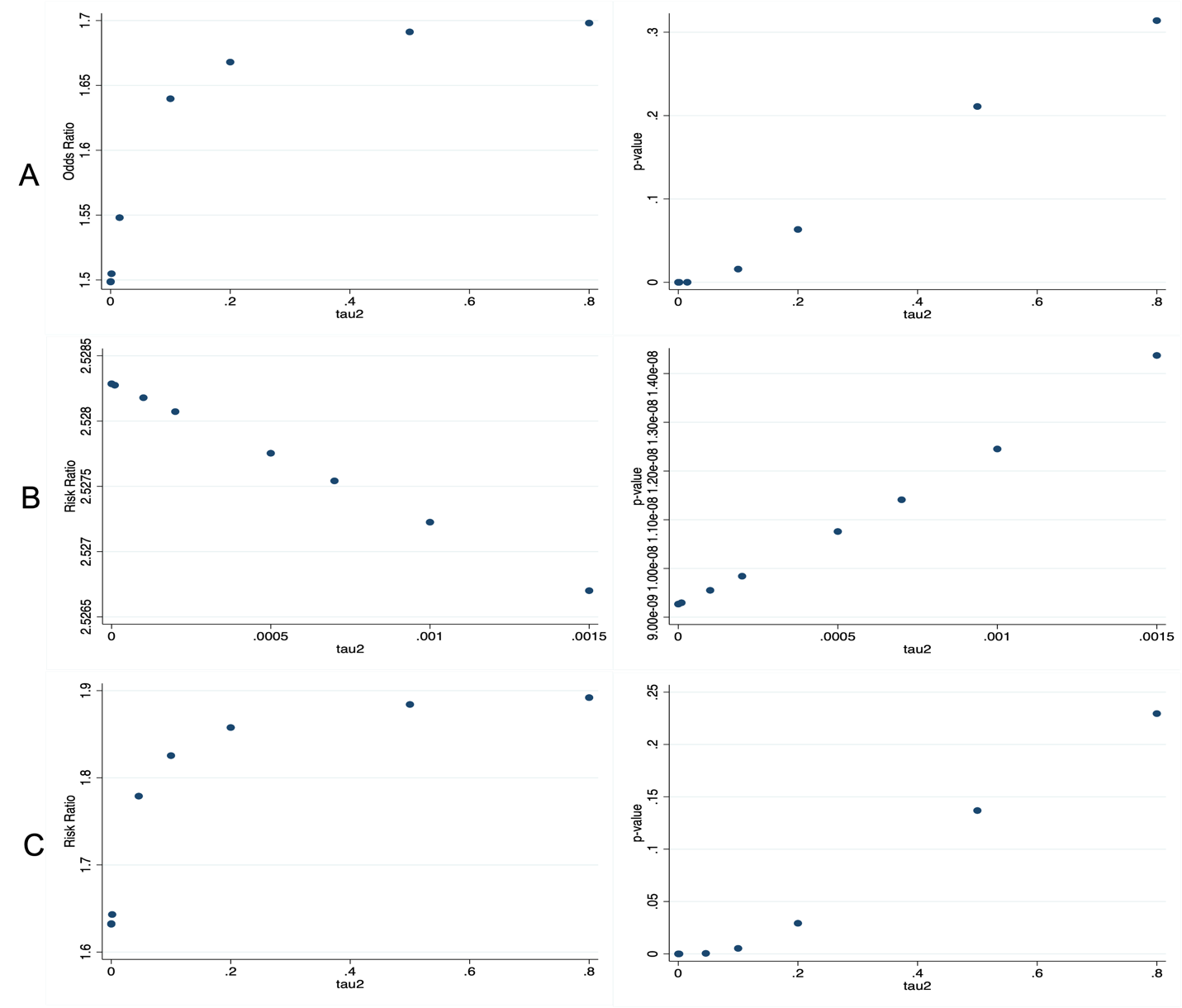
**eAppendix 3**. Sensitivity Analyses of pooled odds ratios and relative risk ratios. (A) Sensitivity analysis by odds ratio and p-values for ordinal shift in modified Ranking scale (mRS). (B) Sensitivity analysis by risk ratio (RR) and p-value for functional independence (mRS 0-2). (C) Sensitivity analysis by RR and p-value for ambulatory independence mRS (mRS 0-3).

**eAppendix 4**. Publication bias funnel plot for generalized odds ratio for ordinal shift in modified Rankin scale.

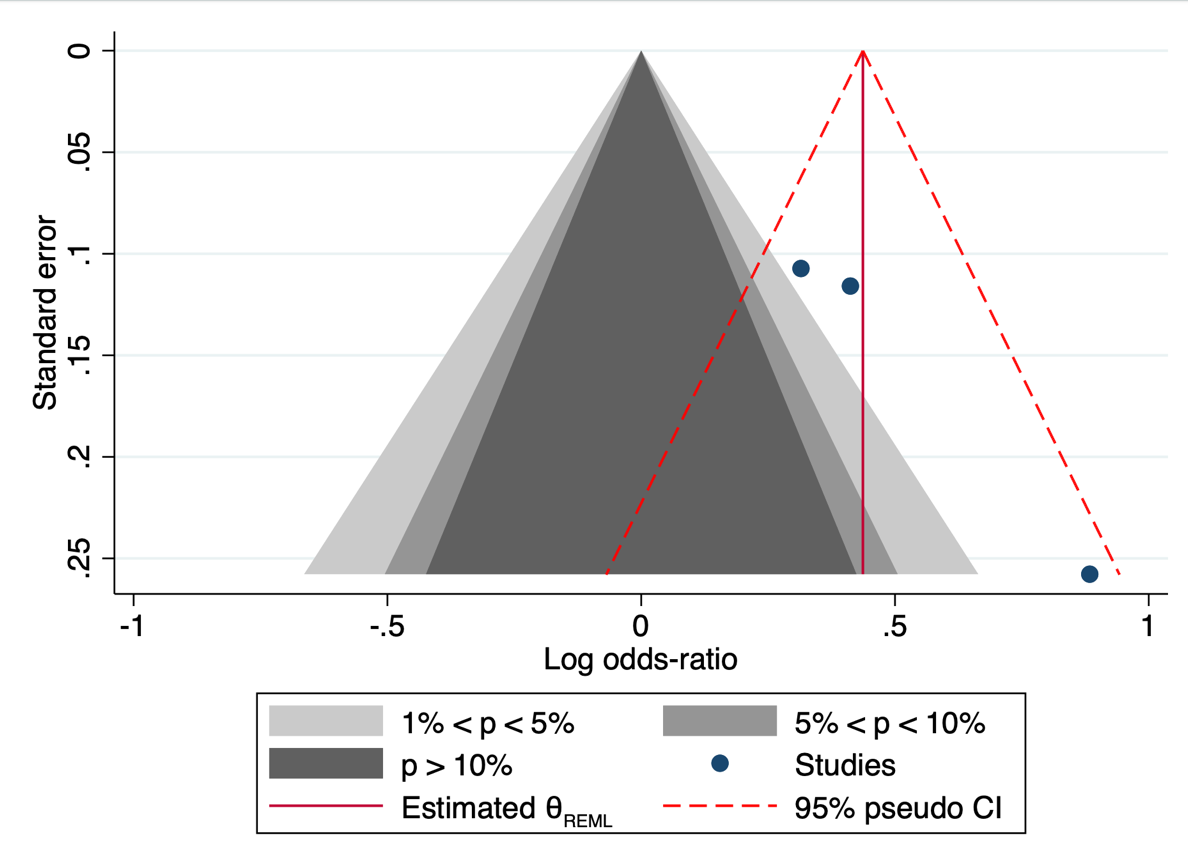

**eAppendix 5**. Grading of Recommendations Assessment, Development, and Evaluation (GRADE) assessment for quality of evidence.

| **Certainty assessment** | | | | | | | **№ of patients** | | **Effect** | | **Certainty** | **Importance** |
| --- | --- | --- | --- | --- | --- | --- | --- | --- | --- | --- | --- | --- |
| **№ of studies** | **Study design** | **Risk of bias** | **Inconsistency** | **Indirectness** | **Imprecision** | **Other considerations** | **Endovascular therapy** | **medical management** | **Relative (95% CI)** | **Absolute (95% CI)** |  |  |
| **Ordinal shift on the modified Rankin scale at 90 days (follow-up: 90 days; assessed with: modified Rankin scale; Scale from: 0 to 6)** | | | | | | | | | | | | |
| 3 | randomized trials | not serious | not serious | not serious | not serious | none | 509 | 501 | - | odds ratio **0 1.55**  (1.25 higher to 1.91 higher) | ⨁⨁⨁⨁ High |  |
| **Functional independence (follow-up: 90 days; assessed with: modified Rankin scale; Scale from: 0 to 2)** | | | | | | | | | | | | |
| 3 | randomized trials | not serious | not serious | not serious | not serious | none | 509 | 501 | - | odds ratio **0 2.53**  (1.84 higher to 3.47 higher) | ⨁⨁⨁⨁ High |  |
| **Independent Ambulation (follow-up: 90 days; assessed with: modified Rankin scale; Scale from: 0 to 3)** | | | | | | | | | | | | |
| 3 | randomized trials | not serious | not serious | not serious | not serious | none | 509 | 501 | - | odds ratio **0 1.78**  (1.29 higher to 2.46 higher) | ⨁⨁⨁⨁ High |  |
| **Any Intracranial Hemorrhage** | | | | | | | | | | | | |
| 2 | randomized trials | not serious | not serious | not serious | not serious | none | 171/330 (51.8%) | 71/327 (21.7%) | **RR 2.30** (1.51 to 3.49) | **282 more per 1,000** (from 111 more to 541 more) | ⨁⨁⨁⨁ High |  |
| **Symptomatic Intracranial Hemorrhage** | | | | | | | | | | | | |
| 3 | randomized trials | not serious | not serious | not serious | not serious | none | 24/509 (4.7%) | 13/501 (2.6%) | **RR 1.85** (0.94 to 3.63) | **22 more per 1,000** (from 2 fewer to 68 more) | ⨁⨁⨁⨁ High |  |
| **Death (follow-up: 90 days)** | | | | | | | | | | | | |
| 3 | randomized trials | not serious | not serious | not serious | not serious | none | 136/509 (26.7%) | 140/501 (27.9%) | **RR 0.95** (0.78 to 1.16) | **14 fewer per 1,000** (from 61 fewer to 45 more) | ⨁⨁⨁⨁ High |  |
| **Decompressive Hemicraniectomy** | | | | | | | | | | | | |
| 2 | randomized trials | not serious | not serious | not serious | not serious | none | 27/330 (8.2%) | 22/327 (6.7%) | **RR 1.22** (0.44 to 3.40) | **15 more per 1,000** (from 38 fewer to 161 more) | ⨁⨁⨁⨁ High |  |

**CI:** confidence interval; **RR:** risk ratio
